# Low levels of monkeypox virus neutralizing antibodies after MVA-BN vaccination in healthy individuals

**DOI:** 10.1101/2022.08.31.22279414

**Authors:** Luca M. Zaeck, Mart M. Lamers, Babs E. Verstrepen, Theo M. Bestebroer, Martin E. van Royen, Hannelore Götz, Marc C. Shamier, Leanne P.M. van Leeuwen, Katharina S. Schmitz, Kimberley Alblas, Suzanne van Efferen, Susanne Bogers, Sandra Scherbeijn, Guus F. Rimmelzwaan, Eric C.M. van Gorp, Marion P.G. Koopmans, Bart L. Haagmans, Corine H. GeurtsvanKessel, Rory D. de Vries

## Abstract

In July 2022, the ongoing monkeypox (MPX) outbreak was declared a public health emergency of international concern by the World Health Organization. Modified vaccinia virus Ankara (MVA-BN, also known as Imvamune, Jynneos, or Imvanex) is a 3^rd^ generation smallpox vaccine that was generated by serial passaging of the more pathogenic parental vaccinia virus (VACV), and is authorized as a vaccine against MPX in humans in a two-shot regimen. Up to now, there is a lack of data that demonstrate MPX virus (MPXV)-neutralizing antibodies in vaccinated individuals and vaccine efficacy data against MPXV infection. Here, we measure MVA-, VACV-, and MPXV-reactive binding and neutralizing antibodies with validated in-house assays in cohorts of historically smallpox-vaccinated, MPXV PCR-positive, and recently MVA-BN-vaccinated individuals. We show that MPXV neutralizing antibodies were detected across all cohorts in individuals with MPXV exposure as well as those who received historic (VACV) vaccination. However, a primary MVA-BN immunization series in non-primed individuals yields relatively low levels of MPXV neutralizing antibodies. As the role of MPXV neutralizing antibodies for protection against disease and transmissibility is currently unclear and no correlate of protection against MPXV infection has been identified yet, this raises the question how well vaccinated individuals are protected. Dose-sparing leads to lower antibody levels, whereas a third MVA vaccination further boosts the antibody response. Cohort studies following vaccinated individuals are necessary to further assess vaccine efficacy in risk populations and determine correlates of protection for this emerging pathogen.

## Introduction

Monkeypox virus (MPXV) belongs to the Orthopoxvirus genus of the Poxviridae family of large double-stranded DNA viruses, and causes a zoonotic disease known as monkeypox (MPX). In May 2022, MPX was identified in several countries in which MPX cases had not been reported previously, after which MPXV rapidly spread in Europe and the United States among individuals who had not traveled to endemic areas.^1^ On July 23, 2022, this ongoing MPX outbreak was declared a public health emergency of international concern (PHEIC) by the World Health Organization (WHO).^2^

MPXV is closely related to variola virus (VARV), the causative agent of smallpox. Smallpox was eradicated by the use of different attenuated poxvirus vaccines combined with active case finding, isolation, and quarantine measures. The 1st and 2nd generation smallpox vaccines contained infectious vaccinia virus (VACV) grown either in the skin of live animals, the chorioallantoic membrane of eggs, or cell culture. The 3rd generation smallpox vaccine was based on an even further attenuated VACV obtained by serial passage in chicken embryo fibroblasts (CEF), known as modified vaccinia virus Ankara (MVA). Since MVA was developed in the endgame of smallpox eradication (first market authorization was obtained in Germany in 1977),^3^ efficacy against smallpox has been inferred based on the noninferiority of immunogenicity in clinical studies.^4^ A study in the Democratic Republic of the Congo showed that the VACV smallpox vaccine was effective against MPX to a certain extent.^5^ Efficacy data of the 3rd generation MVA smallpox vaccine against MPX in humans is lacking. MVA vaccination afforded protection against severe MPX disease and death in non-human primates by inducing both antibodies and T-cells, but sterile immunity was not achieved and some skin lesions remained.^6-8^ Partly because of this evidence for protection from severe disease, MVA-BN (named after manufacturer Bavarian Nordic) was licensed as a vaccine against MPX in humans in Canada (known as Imvamune) and the United States (known as Jynneos), and was recently approved by the European Medicines Agency (EMA) ‘under special circumstances’ (known as Imvanex), despite a lack of efficacy data against human MPXV infection or demonstrable MPXV-neutralizing antibodies in vaccinated individuals.

Randomized trials, ‘test-negative’ studies, and cohort studies are being initiated to better understand MVA-BN efficacy against MPX.^9^ While these studies are underway, it is equally important to better understand the immunogenicity of MVA, especially with regards to MPXV. Assays to measure VACV- and MVA-binding antibodies have been previously developed in ELISA formats using purified intracellular mature virions (IMV) or infected cell lysates.^6,10^ Functional antibody measurements through virus neutralization assays using VACV or MVA expressing reporter proteins have been used for studies assessing the noninferiority of vaccine-induced immunogenicity between new- and old-generation vaccines.^6,10,11^ However, assays to measure MPXV-specific antibodies are lacking. Here, we developed both ELISAs and neutralization assays based on a MPXV isolate from a Dutch patient, to address three crucial questions: (1) are antibodies induced by historic smallpox vaccination cross-reactive with MPXV, (2) do MPXV-infected individuals rapidly mount neutralizing antibody responses, and (3) does MVA-BN vaccination induce MPXV-reactive and neutralizing antibodies?

## Results

### VACV-reactive antibodies detected >70 years after historic smallpox vaccination cross-neutralize MPXV

To determine whether we could detect VACV-reactive IgG antibodies induced by historic smallpox vaccination, we first performed an ELISA with VACV-infected cell lysate and sera selected from the Erasmus MC biobank based on year of birth, and divided over decades ≤1974 (N=56) and >1974 (N=52) (smallpox vaccination was stopped in 1974 in the Netherlands). Sera were all obtained in 2022, meaning that the sera from individuals born prior to 1950 were >70 years post historic smallpox vaccination (median years between sample and birth was 74 years, range 73 – 85 years). VACV-reactive antibodies were frequently detected in sera obtained from individuals born prior to 1974 (42/59, 71%), but hardly in individuals born after 1974 (2/67, 3%) (**Supplemental Table 1, Supplemental Figure 1, Figure 1A**). From the sera tested in ELISA, we randomly selected 21 sera (N=16 and N=5 from individuals born ≤1974 or >1974, respectively; colored symbols in **Figure 1A**) to assess the presence or absence of antibodies capable of neutralizing MPXV. Neutralization of MPXV was exclusively detected in the selection of sera from individuals born prior to 1974 (**Figure 1B**). Sera not capable of neutralizing MPXV were also negative for VACV-reactive antibodies in ELISA.

**Figure 1.**
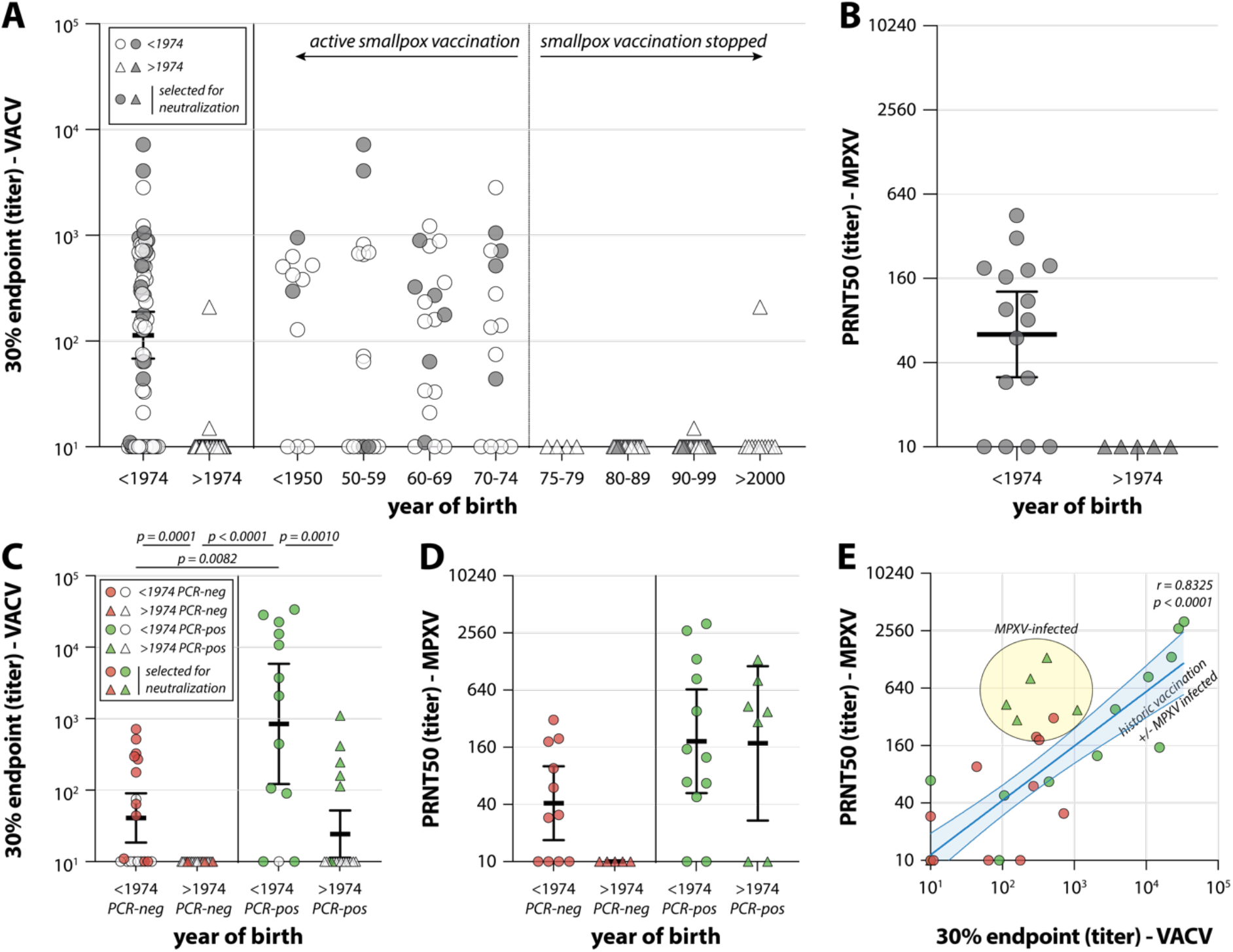
VACV-reactive and MPXV-neutralizing antibodies after historic smallpox vaccination and MPXV infection. (A-B) Detection of poxvirus-specific antibodies in an age-panel of sera: (A) Detection of VACV-reactive antibodies by ELISA against VACV-infected cell lysate. Sera obtained from individuals born in or prior to 1974 (circles), or after 1974 (triangles) are merged on the left side of the graph, and shown per decade on the right side of the graph. Colored symbols reflect sera selected for neutralization assays. (B) Detection of MPXV neutralizing antibodies by plaque reduction neutralization test (PRNT) on a selection of N=21 sera. (C-E) Detection of poxvirus-specific antibodies in a diagnostic panel of sera: (C) Detection of VACV-reactive antibodies by ELISA against VACV-infected cell lysate. Sera were obtained from individuals born in or prior to 1974 (circles), or after 1974 (triangles), who were either PCR-negative (red) or PCR-positive (green). Colored symbols reflect sera selected for neutralization assays. (D) Detection of MPXV-neutralizing antibodies by PRNT on a selection of N=35 sera. (E) Relationship between VACV-reactive binding and MPXV-neutralizing antibodies by correlating the data from panels (C) and (D). 30% endpoint ELISA titers were calculated based on a 5-fold dilution series, after subtraction of OD450 values against a mock-infected cell lysate, and relative to a positive control. The 50% plaque reduction neutralization titer (PRNT50) was calculated on the basis of a 2-fold dilution series relative to an infection control. Lines indicate geometric mean; whiskers indicate 95% confidence interval. A Mann-Whitney U test was performed to compare VACV-reactive endpoint titers (C) and MPXV neutralizing PRNT50 (D) (p<0.0083 considered significant after Bonferroni correction for multiple comparisons), and MPXV neutralizing antibodies were correlated by performing Spearman r analysis (excluding the N=5 sera from MPXV-infected individuals born after 1974).

### MPXV infection induces VACV-reactive antibodies and boosts antibody titers induced by historic smallpox vaccination

To determine whether MPXV infection leads to production of VACV-reactive IgG antibodies, we performed an ELISA against a lysate of VACV-infected cells on diagnostic sera submitted to our laboratory for MPXV RT-PCR. We included sera from both MPXV PCR-negative (N=40, of which N=19 sera from individuals born ≤1974) and PCR-positive individuals (N=32, of which N=13 sera from individuals born ≤1974) (**Supplemental Table 2**). In MPXV PCR-negative individuals, VACV-reactive IgG antibodies were exclusively detected in participants born ≤1974 (N=10/19, 53%), reflecting antibodies induced by historic smallpox vaccination. In MPXV PCR-positive individuals, from whom sera were exclusively obtained in the early symptomatic phase, VACV-reactive IgG antibodies were detected in individuals born ≤1974 (N=10/13, 77%) and >1974 (N=5/19, 26%) (**Supplemental Table 2, Supplemental Figure 2, Figure 1C**). Since we had thus far hardly detected VACV-reactive IgG antibodies in individuals born after 1974, we assume that these were induced by MPXV infection. Antibody responses were more frequently detected in MPXV PCR-positive individuals born ≤1974, and the geometric mean antibody level was significantly higher, compared to historic vaccination by itself (*p*=0.0082, Mann-Whitney U test) or MPXV infection by itself (*p*=0,0010, Mann-Whitney U test), indicative of a rapid recall antibody response induced by MPXV infection (**Figure 1C**, green symbols).

**Figure 2.**
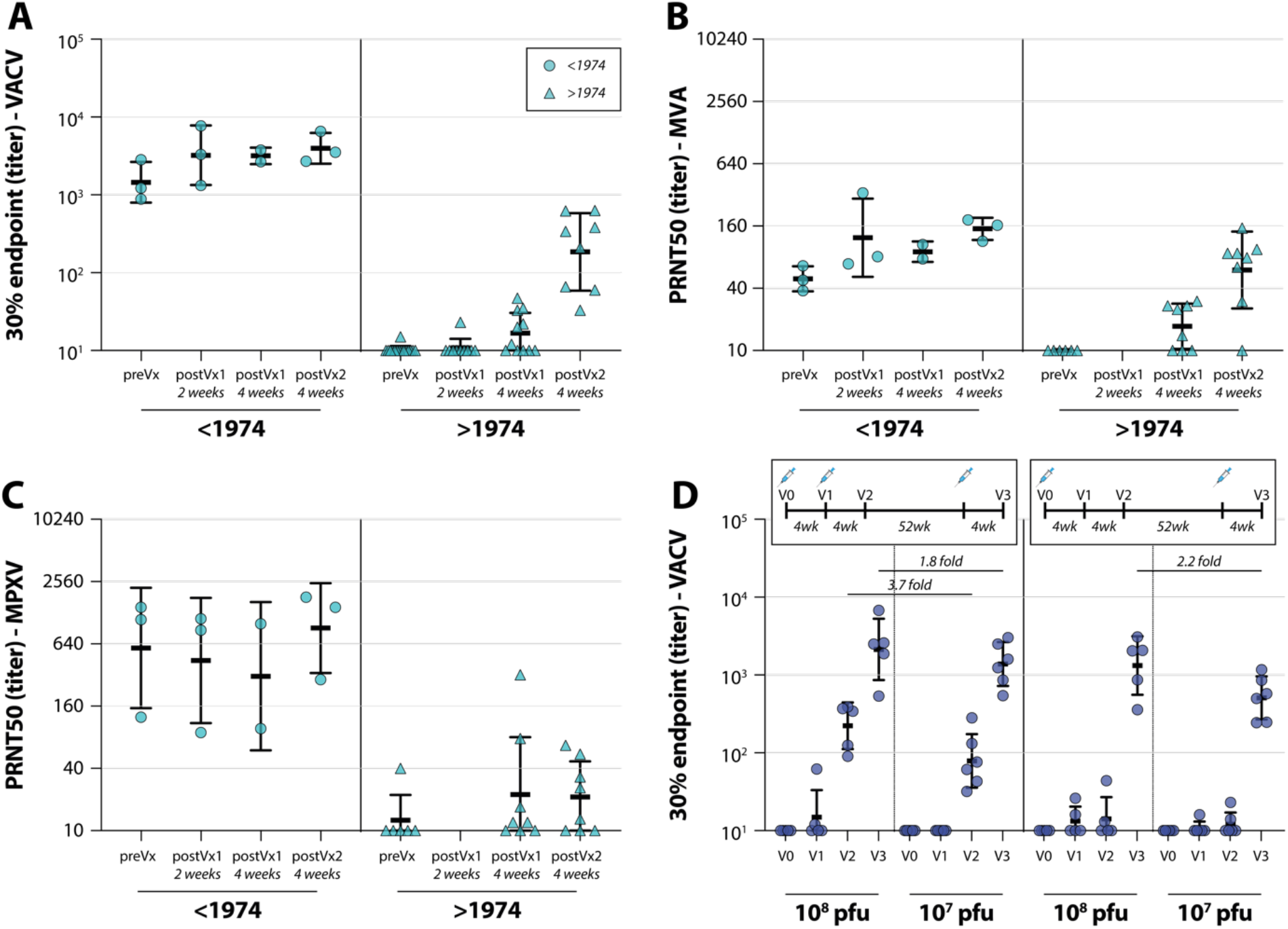
VACV-reactive and MPXV-neutralizing antibodies after Imvanex or MVA-H5 vaccination. (A-C) Detection of poxvirus-specific antibodies in a panel of sera obtained from Imvanex-vaccinated participants. Sera were obtained pre-vaccination and at 3 time points post-vaccination from individuals born in or prior to 1974 (circles), or after 1974 (triangles): (A) Detection of VACV-reactive antibodies by ELISA against VACV-infected cell lysate. (B) Detection of MVA-neutralizing antibodies by PRNT on a serum selection of N=33 sera. (C) Detection of MPXV-neutralizing antibodies by PRNT on a serum selection of N=33 sera. (D) Detection of VACV-reactive antibodies by ELISA against VACV-infected cell lysate. Sera were obtained from participants in an MVA-H5 vaccination trial, who received either a high (10^8^) or low (10^7^) dose regimen, with 2 (right panel) or 3 (left panel) vaccinations. Vaccination regimens are indicated in the legend above the panel. Lines indicate geometric mean; whiskers indicate geometric standard deviation. Fold differences between geometric mean titers after the second or booster vaccination with 10^8^ or 10^7^ pfu are indicated. 30% endpoint ELISA titers were calculated based on a 5-fold dilution series, after subtraction of OD450 values against a mock-infected cell lysate, and relative to a positive control. The 50% plaque reduction neutralization titer (PRNT50) was calculated on the basis of a 2-fold dilution series relative to an infection control.

### Antibodies induced or boosted by MPXV infection neutralize MPXV

We selected 35 sera from MPXV PCR-negative (N=16, of which N=11 were sera from individuals born ≤1974) and PCR-positive individuals (N=19, of which N=12 were sera from individuals born ≤1974) to assess the presence of antibodies capable of neutralizing MPXV (colored symbols in **Figure 1C**). Similar to the ELISA results, virus neutralizing activity in sera from PCR-negative individuals was only observed in individuals born ≤1974, reflective of antibodies induced by historic smallpox vaccination (**Figure 1D**, red symbols). MPXV neutralizing antibodies were also detected in sera from MPXV PCR positive individuals born after 1974. Interestingly, the rapid boosting of VACV-reactive IgG antibodies after MPXV infection of individuals born ≤1974 was not as obvious with respect to MPXV neutralizing antibodies as no significant differences were observed (**Figure 1D**). However, a trend towards higher antibody levels in MPXV PCR-positive individuals born ≤1974 was observed when compared to historic vaccination by itself (*p*=0.0740, Mann-Whitney U test). This effect was better visualized when performing a direct comparison between the VACV-reactive IgG antibodies and MPXV neutralizing antibodies, revealing a good correlation between VACV ELISA and MPXV PRNT50 titers (Spearman correlation r=0.8325, p<0.0001) with the exception of a cluster of sera obtained from exclusively MPXV-infected individuals (**Figure 1E**, green triangles).

### Imvanex vaccination induces VACV-reactive IgG antibodies

To determine whether Imvanex vaccination leads to the production of VACV-reactive IgG antibodies, we performed an ELISA with a lysate of VACV-infected cells and sera obtained pre-vaccination, 2 and 4 weeks after the first vaccination, and 4 weeks after the second vaccination. Participants were vaccinated with the advised dose, 0.5ml with no less than 5 × 10^7^ plaque forming units (pfu). A total of 18 study participants were included (N=3 born ≤1974), of which 11 were followed until the last time-point at the time of writing (**Supplemental Table 3, Supplemental Figure 3**). VACV-reactive IgG antibodies were detected in all sera from study subjects born ≤1974 on all time points. A clear boosting by vaccination was not observed in these three individuals, who already had high binding-antibody levels prior to vaccination (**Figure 2A** left panel, compare to **Figure 1A**). In study subjects born >1974, a gradual increase in binding antibody responses was seen, with detectable antibodies in 1/10 (10%) sera obtained 2 weeks after the first vaccination, 6/11 (55%) sera obtained 4 weeks after the first vaccination, and 8/8 sera obtained 4 weeks after the second vaccination (**Supplemental Table 3** and **Figure 2A**).

### Imvanex vaccination induces low levels of MPXV neutralizing antibodies

In addition to performing an ELISA with a VACV-infected cell lysate, both MVA and MPXV neutralization assays were performed with sera from Imvanex-vaccinated study subjects. Both MVA and MPXV neutralizing antibodies were detected in study subjects born ≤1974 at all timepoints, also prior to Imvanex vaccination (**Figure 2B** and **2C**, left panels). Similar to the VACV-reactive binding antibodies, MVA neutralizing antibodies were induced by vaccination and increased over time in participants born after 1974. Pre-vaccination, 0/6 sera had detectable MVA-neutralizing antibodies, increasing to 5/8 and 8/8 after the first and second vaccination, respectively (**Figure 2B**, right panel). In contrast, MPXV-neutralizing antibodies after vaccination with Imvanex were detected less frequently. Only in fice out of eight sera MPXV neutralizing antibodies were detected 4 weeks after the first, but also the second vaccination. Antibody levels in some vaccinees increased after the second shot, but in general little increase in MPXV neutralization was observed after the second shot (**Figure 2C**, right panel).

### A third MVA vaccination 1 year after completion of the primary regimen significantly boosts VACV-reactive antibodies

To further assess MVA immunogenicity, we performed an ELISA with a lysate of VACV-infected cells and sera from an MVA-H5 influenza vaccination trial.^12^ Sera were obtained from study subjects (who were all born >1974) who received vaccinations following two different regimens: (1) single shot primary vaccination regimen followed by a boost after 1 year, or (2) two shots primary vaccination regimen followed by a boost after 1 year. Additionally, participants received either a high (10^8^ pfu) or low (10^7^ pfu) dose of MVA-H5. Sera were always obtained 4 weeks after vaccination (**Figure 2D**, top legend). We observed similar levels of VACV-reactive antibodies 4 weeks after 2 shots of Imvanex or MVA-H5 (compare **Figure 2D** with **2A**). As for dosing, we found that vaccination with a high dose resulted in more binding antibodies: 3.7-fold higher binding antibody levels when comparing 2 shots of 10^8^ with 2 shots of 10^7^, and 1.8-fold and 2.2-fold higher binding antibody levels when comparing the booster vaccination after a 2 shot or 1 shot primary regimen, respectively (**Supplemental Figure 4, Figure 2D**). Additionally, the second vaccination is important for reaching detectable antibody levels, as individuals in a single shot regimen hardly developed antibody responses 4 and 8 weeks after vaccination. Finally, a third vaccination given after 1 year boosted the binding antibody levels in all dosing groups (10- and 19-fold in the 2 shot regimen for 10^8^ or 10^7^ dose respectively, over 50-fold for both doses in the 1 shot regimen) (**Supplemental Table 4** and **Figure 2D**).

### ELISAs with VACV-infected cell lysate and PRNTs against MPXV grown on Calu-3 cells are most sensitive for antibody detection

Thus far, we assessed the presence of binding and neutralizing antibodies in sera with an ELISA against a VACV-infected cell lysate and PRNTs with infectious MPXV grown on Calu-3 cells. We additionally developed an ELISA with a MPXV-infected cell lysate, and directly compared the 30% endpoint titers obtained from either the VACV or MPXV ELISA for the age-panel, diagnostic panel, and Imvanex panel of sera (**Supplemental Figure 5A**, original data in **Figure 1A, 1C** and **1F**). The ELISA with MPXV-infected cell lysate appeared less sensitive, and only detected antibodies if the 30% endpoint titer against VACV was >1000. Additionally, we compared PRNT50 titers against infectious MPXV grown on either Calu-3 or Vero cells for the age-panel and diagnostic panel of sera (**Supplemental Figure 5B**). The PRNT against Vero-grown MPXV was considerably less sensitive, predominantly detecting neutralizing antibodies at PRNT50 values of >640 against Calu-3-grown MPXV.

## Discussion

Here, we measured MVA-, VACV- and MPXV-reactive binding and neutralizing antibodies in cohorts of historic smallpox-vaccinated, MPXV PCR-positive, MVA-BN-vaccinated, and MVA-H5-vaccinated individuals. For the development and validation of novel assays, an MPXV isolate obtained during the ongoing outbreak was used. MPXV neutralizing antibodies were detected across all cohorts in individuals with MPXV exposure as well as those who received historic (VACV) or recent (MVA-BN or MVA-H5) poxvirus vaccination. A clear boosting effect of MPXV infection on binding antibody levels was observed in previously vaccinated individuals. Simultaneously, this boosting effect was not as apparent for individuals who had received a recent MVA-BN vaccination after historic smallpox vaccination, although there was a trend towards significance. Strikingly, relatively low binding antibody levels were observed after two shots of MVA-BN in individuals without pre-existing poxvirus-specific immune responses, with poor neutralizing capacity.

Although little is known about the antigenic similarities between poxviruses, MVA-BN immunogenicity has thus far only been assessed by measuring MVA- and VACV-specific antibodies;^4^ cross-reactivity with other poxviruses is often assumed. We argue that for use of this vaccine during an ongoing MPXV outbreak, it is essential to measure functionality of vaccine-induced antibodies against the currently circulating MPXV strain. To assess antigenic similarities between poxviruses and select the most appropriate serological assays for studying MPXV-reactive immune responses, we compared binding and neutralizing activity of sera from infected and/or vaccinated individuals against VACV, MVA, and/or MPXV. Measuring VACV-reactive binding antibodies by ELISA proved sensitive, as serological responses were detected in the majority of sera from participants born prior to 1974, and in the majority of recent vaccinees. Since we did not have access to historic vaccination records, we could not confirm whether people born prior to 1974 were vaccinated against smallpox, how many shots they received, and which vaccine was used. Measuring MPXV-reactive binding antibodies with an in-house developed ELISA proved less sensitive.

Because the currently used vaccine is based on MVA, properly assessing vaccine immunogenicity should involve measuring both MVA-specific and MPXV cross-reactive neutralizing antibodies. Interestingly, only limited correlation was observed between MVA and MPXV neutralization in MVA-BN-vaccinated individuals, indicative of antigenic differences between these poxviruses. Depending on the research question, this suggests that measuring a combination of both VACV-reactive antibodies, and MVA and MPXV neutralizing antibodies may be required to study vaccine immunogenicity.

The evidence for cross-protection afforded by VACV or MVA vaccination against MPX is inferred from animal experiments and from observational studies conducted during the period of enhanced surveillance in the endgame of smallpox eradication. In those studies, partial clinical protection was observed. In our study, the individuals born prior to 1974 still had detectable antibodies that neutralized MPXV, yet current epidemiological data suggest limited protection from infection in this age group (diagnostic observations). The primary MVA immunization series in non-primed individuals yielded relatively low levels of neutralizing antibodies, raising the question whether vaccinated individuals are now protected, and what the correlates of protection against MPXV infection are. In non-human primates, depletion studies underlined that antibodies do play an important role against lethal intravenous MPXV challenge,^13^ although both virus-specific antibodies and T-cells were induced by MVA-BN vaccination. At this moment it is unclear what the relatively low MPXV neutralizing titers mean for protection against disease and transmissibility. Finally, by using a serum set from a previously performed MVA-H5 clinical trial, we showed that VACV-reactive antibodies can be further boosted with an additional shot of MVA. This same trial indicates that dose-sparing (10^7^ instead of 10^8^ pfu) has a negative effect on the serological outcome of vaccination. Cohort studies following vaccinated individuals and including biological sampling are necessary to further assess vaccine efficacy in risk populations and determine correlates of protection for this emerging pathogen.

## Materials & Methods

### Cell culture

CEF were isolated from 11-day-old chicken embryos (Drost Loosdrecht BV) and passaged once before use. CEF were cultured in virus production serum-free medium (VP-SFM; Gibco) containing penicillin and streptomycin (P/S). Baby hamster kidney 21 (BHK-21) cells were cultured in Dulbecco’s modified Eagle medium (DMEM; Lonza) supplemented with 10% FBS, 20 mM HEPES, 0.1% CHNaO3, 0.1 mM nonessential amino acids (NEAA; Lonza), and P/S/L-glutamine (P/S/G). HeLa cells were cultured in DMEM supplemented with 10% FBS, 20 mM HEPES, 0.1% CHNaO3, and P/S/G. Vero cells were cultured in DMEM (Capricorn Scientific) supplemented with 10% FBS, 20 mM HEPES, and P/S/G. Calu-3 cells were cultured in Opti-MEM + GlutaMAX (Gibco) supplemented with 10% FBS. All cell lines were grown at 37°C in a humidified CO_2_ incubator.

### Generation of rMVA-GFP and VACV-Elstree virus

rMVA-GFP was generated by homologous recombination as described previously.^14^ MVA clonal isolate F6 served as the parental virus for generating rMVA-GFP.^15^ Vector plasmid pG06-P11-GFP was used to direct the insertion of GFP under the transcriptional control of the natural vaccinia virus (VACV) late promoter P11 into deletion III site of the MVA genome. Virus stocks were generated in CEF, purified by ultracentrifugation through 36% sucrose, and reconstituted in a 120 mM NaCl 10 mM Tris-HCl buffer (pH 7.4). Titer of the rMVA-GFP stock was initially determined by plaque assay on CEF, and was confirmed for PRNT by titration on Vero cells. The stock was validated by PCR, sequencing, and transgene expression in various cell types. The VACV strain Elstree was grown in HeLa cells to serve as ELISA antigen.

### Generation of MPXV

MPXV was isolated from a swab taken from a typical pox lesion of an MPX-positive Dutch patient by inoculating Vero cells. The isolate was designated as MPXV_2022_NL001 and is available through the European Virus Archive (EVAg; Ref-SKU: 010V-04721). Virus stocks were propagated to passage 3 by inoculating 70-90% confluent Vero and Calu-3 cultures grown in T175 flasks at an MOI of 0.1 in Advanced DMEM/F12 (Gibco) supplemented with 10 mM HEPES, 1X GlutaMAX and 1X primocin (AdDF+++). After 4 days, when at least 50% of the surface area in the cultures consisted of visible plaques, cells were harvested using a cell scraper and centrifuged at 2000 x g for 2 min. Cell pellets were resuspended in 500 µl Opti-MEM + GlutaMAX and pipetted up and down to mix 10 times using a P1000 tip. Cell suspensions were lysed by freeze-thawing 3 times in a dry-ice ethanol bath, after which lysates were mixed by pipetting up and down 50 times using a P1000 tip. 10 ml Opti-MEM + GlutaMAX were added to the lysates, which were then cleared by centrifugation at 2000 x g for 5 min. Cleared lysates were filtered through a low protein binding 0.45 µm syringe filter (Millipore), aliquoted, and frozen at −80°C. All work with infectious MPXV was performed in a Class II Biosafety Cabinet under BSL-3 conditions.

### MPXV stock titrations

Stock titers were determined by preparing 10-fold serial dilutions in AdDF+++. One hundred μl of each dilution was added to Vero cells in a 96-well plate and incubated for 16 hours in a humidified CO_2_ incubator at 37°C. Next, cells were fixed in 10% neutral buffered formalin (NBF) for 30 min and permeabilized in 70% ethanol (submerging the entire plate). Cells were then washed in PBS, blocked in 0.6% BSA (Sigma) and 0.1% Triton X-100 (Sigma-Aldrich) in PBS for 30 min, and stained overnight at room temperature with rabbit-anti-VACV-FITC (Abbexa) at a 1:1000 dilution. After washing in PBS, plates were scanned on the Amersham Typhoon Biomolecular Imager (channel Cy2; resolution 10 mm; GE Healthcare). Numbers of infected cells were quantified using ImageQuant TL 8.2 (GE Healthcare).

### Detection of VACV- or MPXV-specific IgG antibodies by ELISA

For the detection of VACV or MPXV-specific antibodies, HeLa or Vero cells were mock-treated or infected with VACV-Elstree or MPXV_2022_NL001 at MOI 1 and 0.1, respectively, and harvested in 1% Triton X-100 in PBS supplemented with mini cOmplete EDTA free protease inhibitor (Roche) when complete cytopathic effect was observed. All work with the MPXV lysate was performed under BSL-3 conditions. For ELISA, high-binding 96-well plates (Corning) were coated for 1 h at 37 °C with the diluted cell lysates (VACV coating: 1:500; MPXV coating: 1:100) in PBS. The coating concentrations were optimized in coating titration experiments. Coated plates were washed 5 times with PBS supplemented with 0.05% Tween-20 (PBST, Merck) and subsequently blocked for 1 h at 37 °C with blocking buffer (PBST + 2% skim milk powder (w/v, Merck)). Fivefold dilution series of sera in blocking buffer (starting dilution 1:10), were transferred to the lysate-coated plates and incubated overnight at 4 °C. Plates were washed with five times with PBST and incubated for 1 h at 37 °C with HRP-conjugated goat-anti-human IgG (Dako). Afterwards, plates were again washed five times with PBST and incubated for about 15 min with 100 µl TMB peroxidase substrate (SeraCare/KPL) after which the reaction was stopped with an equal volume of 0.5 N H2SO4 (Merck). Absorbance was measured at 450 nm using a Tecan Infinite F200 or an Anthos 2001 microplate reader and corrected for absorbance at 620 nm. OD450 values obtained with mock-infected cell lysates were subtracted from the OD450 value obtained with the VACV/MPXV-infected cell lysates to determine a net OD450 response. A positive control serum was included on every ELISA plate, generating a min-to-max OD450 S-curve. OD450 values generated by dilution series per sample were transformed to this control S-curve, and 30% endpoint titers were calculated.

### Detection of MVA/MPXV-specific neutralizing antibodies by PRNT

Vero cells were seeded one day prior to the experiment in 96-well plates (Greiner Bio-One) at a density of 20,000 cells per well. Sera were heat inactivated for 1 hour at 60°C and subsequently 2-fold serially diluted in AdDF+++ before 1500 PFU of MVA-GFP or 400 PFU of MPXV_2022_NL001 in 60 μl were added per well. The final serum dilution in the first column was 1:20. The virus-serum mix was then incubated for 1 h at 37°C before 100 ul of it were added to the Vero cells. The cells were incubated for 24 h (MVA-GFP) or 16 h (MPXV) at 37°C and 5% CO_2_ before fixing in 4% paraformaldehyde for 10 min (MVA-GFP) or in 10% NBF for 30 min (MPXV). MPXV-infected samples were furthermore permeabilized in 70% ethanol, followed by a wash with PBS and blocking in 0.6% BSA and 0.1% Triton X-100 in PBS for 30 min before being stained overnight at room temperature with rabbit-anti-VACV-FITC (Abbexa) at a 1:1000 dilution. Both MVA and MPXV neutralization assays were washed with PBS prior to nuclear staining with Hoechst33342 (Thermo Fisher). Cells were imaged using the Opera Phenix spinning disk confocal HCS system (Perkin Elmer) equipped with a 10x air objective (NA 0.3) and 405 nm and 488 nm solid state lasers. Hoechst and GFP/FITC were detected using 435-480 nm and 500-550 nm emission filters, respectively. Nine fields per well were imaged covering approximately 50% of the individual wells. The number of infected cells was quantified using the Harmony software (version 4.9, Perkin Elmer). The dilution that would yield 50% reduction of plaques compared with the infection control was estimated by determining the proportionate distance between two dilutions from which an endpoint titer was calculated. When no neutralization was observed, the PRNT50 was given a value of 10.

### Serum samples and ethics statement

Serum samples from N=238 participants were included in this study divided over four different cohorts (**Supplemental Table 1-4, Supplemental Figure 1-4**). (1) Age-panel cohort. To validate the assays, an anonymized age-panel cohort was used. For this purpose, samples were randomly selected from the diagnostic serum bank at Erasmus MC based on year of birth, excluding immunocompromised patients. In total, N=126 sera obtained in 2022 were included in this cohort (N=59 born in or prior to 1974, N=67 born after 1974. The Erasmus MC Medical Ethics Committee waived ethical approval for use of these anonymized samples. (2) Diagnostic cohort. In the Netherlands, diagnostic serum samples were drawn in addition to swab samples for PCR testing from patients suspected for MPXV infection. In total, N=72 anonymized diagnostic sera were included in this study, subdivided in PCR-negative and PCR-positive patients, born either in or prior to 1974 or after 1974. The Erasmus MC Medical Ethics Committee waived ethical approval for use of these anonymized samples. (3) Imvanex cohort. Serum samples were obtained from healthcare workers (HCW) who received Imvanex vaccination for safety reasons as employees of a BLS-3 laboratory. Samples were collected on a biobanking study protocol. The Erasmus MC Medical Ethics Committee gave ethical approval for this work performed as part of the COVA study (ethical permit MEC-2014-398). Longitudinal samples were obtained pre-vaccination, 2 and 4 weeks after the first vaccination, and 4 weeks after the second vaccination. Participants were vaccinated with the prescribed dose, 0.5ml with no less than 5 × 10^7^ plaque forming units (pfu). A total of 18 participants were included (N=3 born ≤1974), of which 11 were followed until the last time-point at the time of writing. (4) MVA-H5 cohort. The fourth serum panel consisted of samples that were obtained from participants as part of a past clinical phase I vaccination trial with MVA-H5 in two different regimens (**Figure 2D**), either with a low (10^7^) or high dose (10^8^).^12^ Longitudinal samples were obtained pre-vaccination, 4 weeks after the first vaccination, 4 weeks after the second vaccination or 8 weeks after the first vaccination, and 4 weeks after the booster vaccination after 1 year. The Erasmus MC Medical Ethics Committee gave ethical approval for this work performed in the FluVec-H5 study (ethical permit METC NL37002.000.12, Dutch Trial Registry NTR3401). A total of 22 participants were included here, all born after 1974.

### Data acquisition and statistical analysis

All samples from each respective experimental panel (age-panel [1], diagnostic panel [2], Imvanex-vaccinated panel [3], and MVA-H5-vaccinated panel [4]) were analyzed simultaneously per assay to counteract batch effects. Moreover, assays were standardized on a reference control that was included on every individual assay plate. For ELISAs with VACV-infected cell lysate, this was a pool of two high-titer sera from individuals who received both historic smallpox and recent Imvanex vaccination. For ELISAs using MXPV-infected cell lysates, this was an individual who received historic smallpox vaccination and had recent MPXV exposure. Neutralization assays included an infection control. The reference samples on every plate were used to generate S-curves (**Supplemental Figure 1-4**) to which the test samples were transformed to calculate 30% endpoint titers (ELISA) or PRNT50 values (neutralization assays) as described above. A statistical comparison within VACV-reactive 30% endpoint titers (**Figure 1C**) and MPXV-neutralizing PRNT50 (**Figure 1D**) was performed using a Mann-Whitney U test. Correlation of VACV-reactive 30% endpoints titers and MPXV-neutralizing PRNT50 was evaluated by Spearman *R*. A *p*-value below 0.0083 was considered significant after Bonferroni correction for multiple comparisons. Statistical evaluation was done with GraphPad Prism v9.02.

## Data Availability

All data produced in the present study are available upon reasonable request to the authors.

## Acknowledgements

This work was partially funded from European Union’s Horizon 2020 Grant ECRAID (grant no 965313), in preparation for possible clinical studies and trials following the decision to activate ECRAID to outbreak mode 2. Sequencing work was supported by European Union’s Horizon 2020 Grant VEO (Grant No. 874735).

## SUPPLEMENTAL MATERIAL

### Supplemental Figures

**Supplemental Figure 1.**
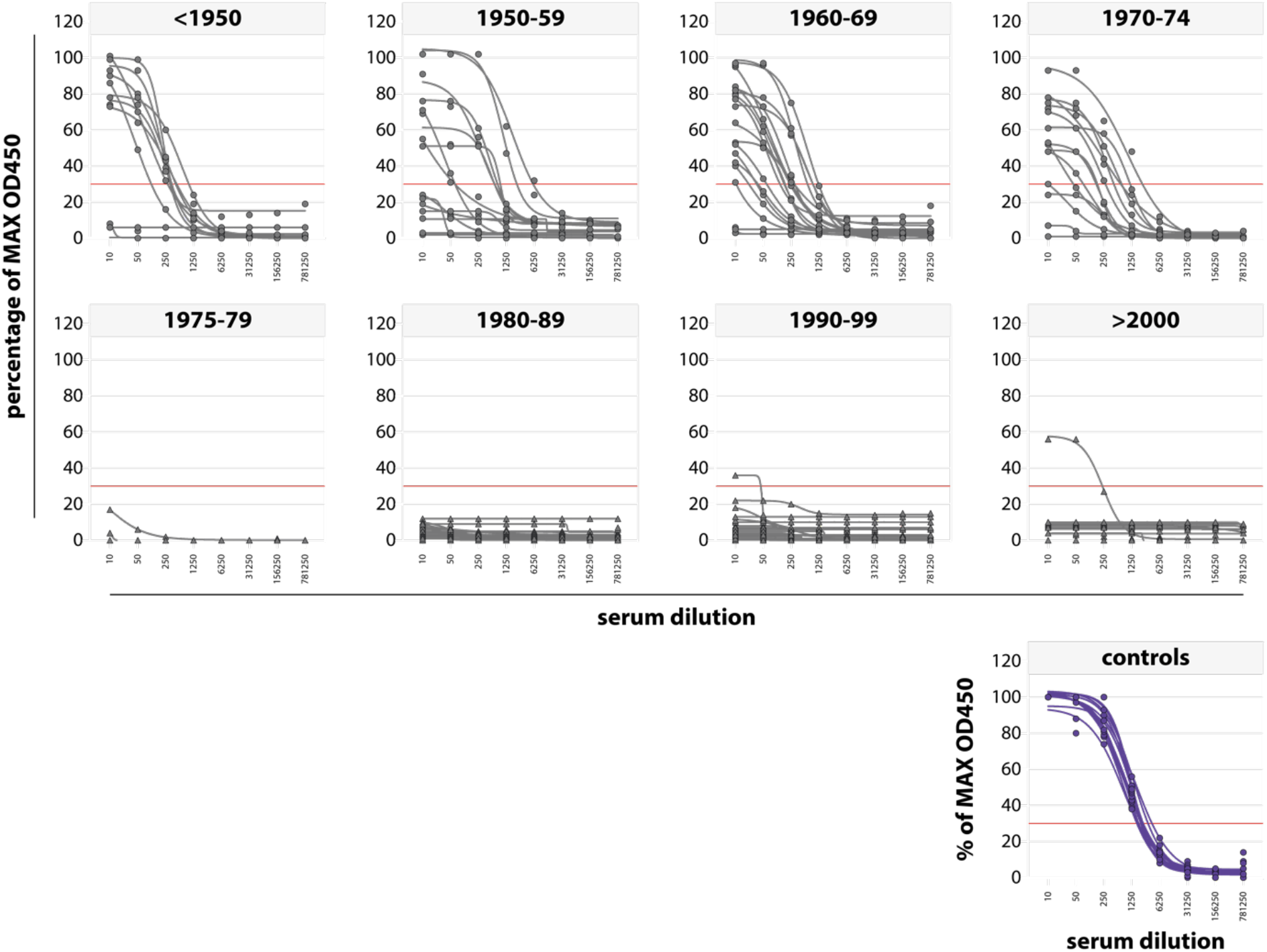
Normalized S-curves for determining the 30% OD450 ELISA endpoint in age-panel sera. Sera from the age-panel were 5-fold diluted starting at a dilution of 1:10 and added to plates coated with a VACV-infected cell lysate. A positive reference control was included on every plate to generate an S-curve with minimum (0%) and maximum (100%) OD450 values. All sample OD450 values were normalized to the positive control on the respective plate and plotted as log (inhibitor) vs response curve with four parameter variable slope using GraphPad Prism 9.02. A 30% endpoint dilution was estimated by calculating the proportionate distance between two dilutions, from which a titer was determined. Symbols indicate individual donors, gray lines represent samples from the age-panel, purple lines represent the controls from the different plates. 30% endpoint titers are shown in **Figure 1A**. All samples from this serum panel were run in the same experiment, avoiding any possible batch effects.

**Supplemental Figure 2.**
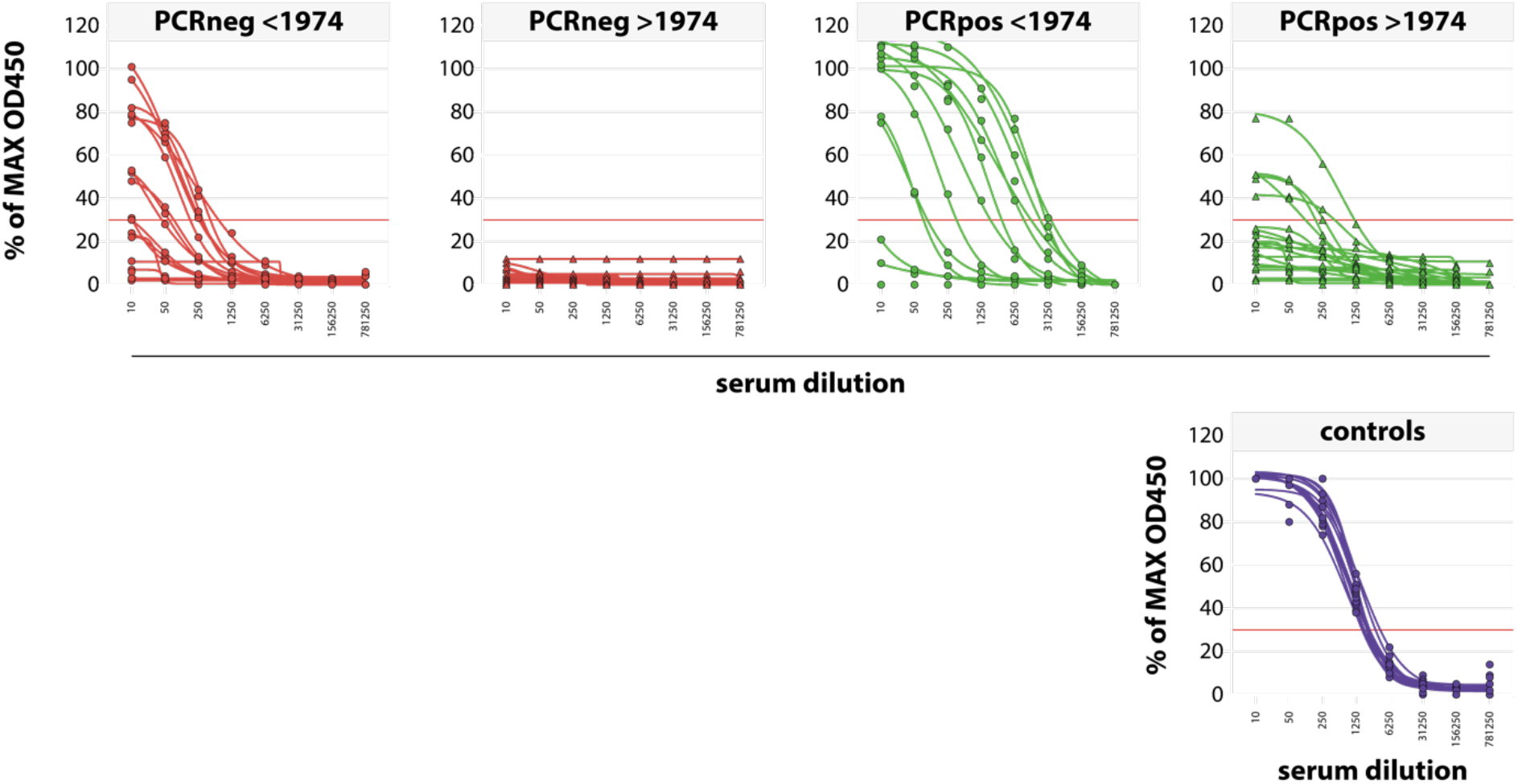
Normalized S-curves for determining the 30% OD450 ELISA endpoint in diagnostic sera. Sera from the diagnostic panel were 5-fold diluted starting at a dilution of 1:10 and added to plates coated with a VACV-infected cell lysate. A positive reference control was included on every plate to generate an S-curve with minimum (0%) and maximum (100%) OD450 values. All’ sample’ OD450 values were normalized to the positive control on the respective plate and plotted as log (inhibitor) vs response curve with four parameter variable slope using GraphPad Prism 9.02. A 30% endpoint dilution was estimated by calculating the proportionate distance between two dilutions, from which a titer was determined. Symbols indicate individual donors (circle: born prior to 1974, triangle born after 1974), red lines represent sera from PCR-negative donors, green lines represent sera from PCR-positive donors, and purple lines represent the controls from the different plates. 30% endpoint titers are shown in **Figure 1C**. All samples from this serum panel were run in the same experiment, avoiding any possible batch effects.

**Supplemental Figure 3.**
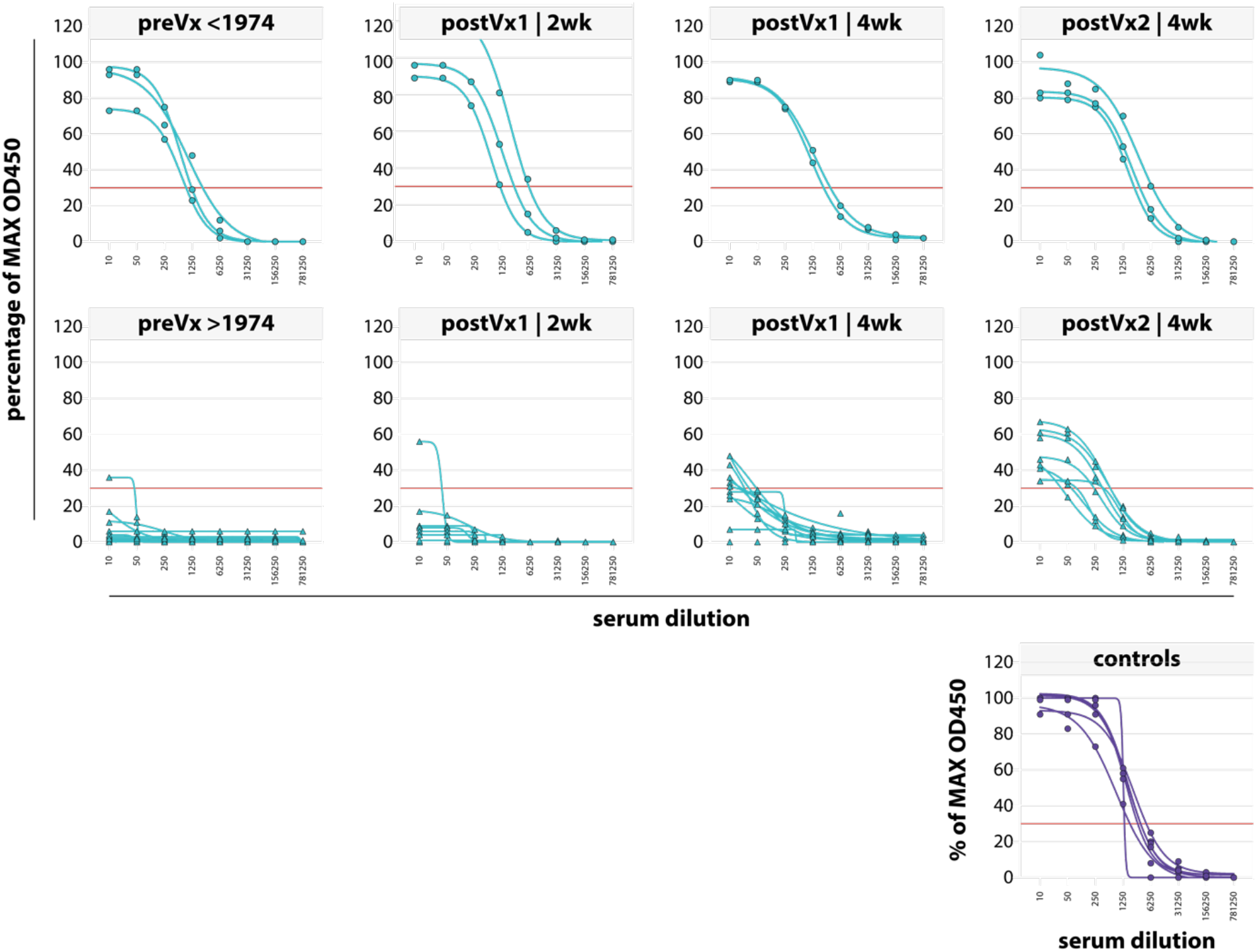
Normalized S-curves for determining the 30% OD450 ELISA endpoint in Imvanex sera. Sera from the Imvanex panel were 5-fold diluted starting at a dilution of 1:10 and added to plates coated with a VACV-infected cell lysate. A positive reference control was included on every plate to generate an S-curve with minimum (0%) and maximum (100%) OD450 values. All’ sample’ OD450 values were normalized to the positive control on the respective plate and plotted as log (inhibitor) vs response curve with four parameter variable slope using GraphPad Prism 9.02. A 30% endpoint dilution was estimated by calculating the proportionate distance between two dilutions, from which a titer was determined. Symbols indicate individual donors (circle: born prior to 1974, triangle born after 1974), cyan lines represent sera from Imvanex-vaccinated donors, purple lines represent the controls from the different plates. 30% endpoint titers are shown in **Figure 2A**. All samples from this serum panel were run in the same experiment, avoiding any possible batch effects.

**Supplemental Figure 4.**
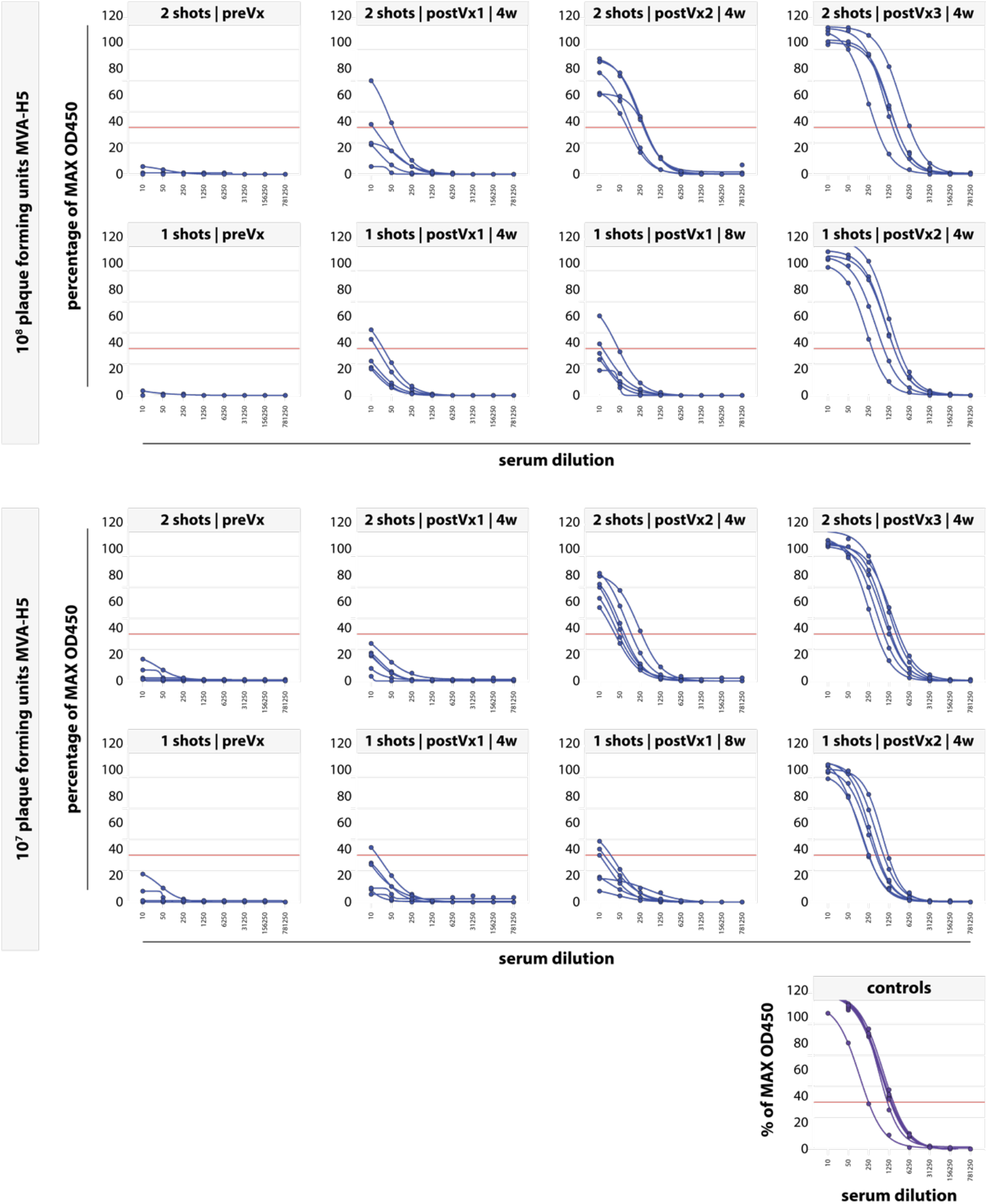
Normalized S-curves for determining the 30% OD450 ELISA endpoint in MVA-H5 sera. Sera from the MVA-H5 panel were 5-fold diluted starting at a dilution of 1:10 and added to plates coated with a VACV-infected cell lysate. A positive reference control was included on every plate to generate an S-curve with minimum (0%) and maximum (100%) OD450 values. All’ sample’ OD450 values were normalized to the positive control on the respective plate and plotted as log (inhibitor) vs response curve with four parameter variable slope using GraphPad Prism 9.02. A 30% endpoint dilution was estimated by calculating the proportionate distance between two dilutions, from which a titer was determined. Symbols indicate individual donors, blue lines represent sera from MVA-H5-vaccinated donors, purple lines represent the controls from the different plates. 30% endpoint titers are shown in **Figure 2D**. All samples from this serum panel were run in the same experiment, avoiding any possible batch effects.

**Supplemental Figure 5.**
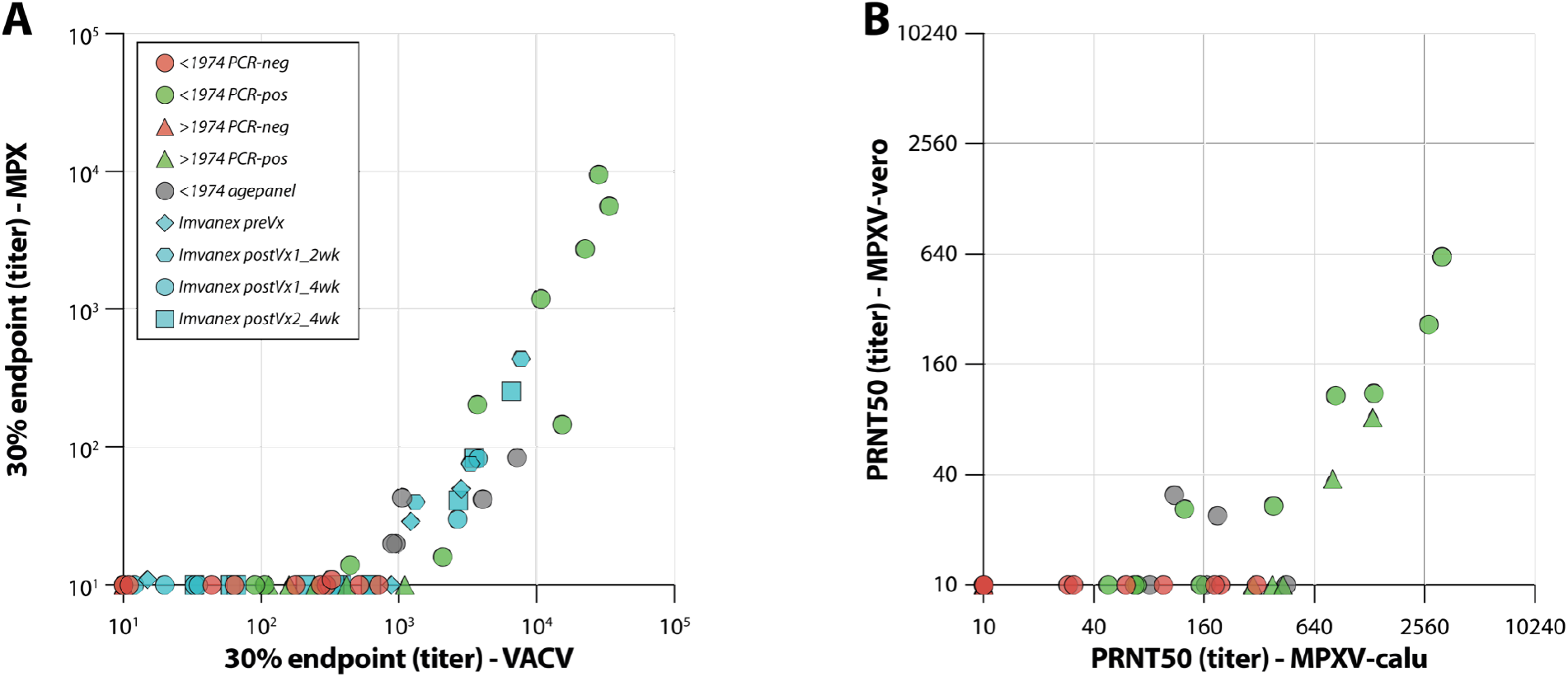
Correlation between ELISAs with VACV and MPXV antigen, and neutralization assays with MPXV grown on Vero or Calu-3 cells. (A) ELISAs were performed on the age-panel, diagnostic panel, and Imvanex panel of sera against a VACV-infected or MPXV-infected cell lysate. 30% endpoint ELISA titers were calculated based on a 5-fold dilution series, after subtraction of OD450 values against a mock-infected cell lysate, and relative to a positive control. (B) PRNTs were performed with an MPXV stock grown on either Calu-3 or Vero cells. The 50% plaque reduction neutralization titer (PRNT50) was calculated on the basis of a 2-fold dilution series relative to an infection control. Sera were obtained from individuals born in or prior to (circles), or after (triangles) 1974, who were either PCR-negative (red and gray) or PCR-positive (green).

### Supplemental Tables

**Supplemental Table 1.**
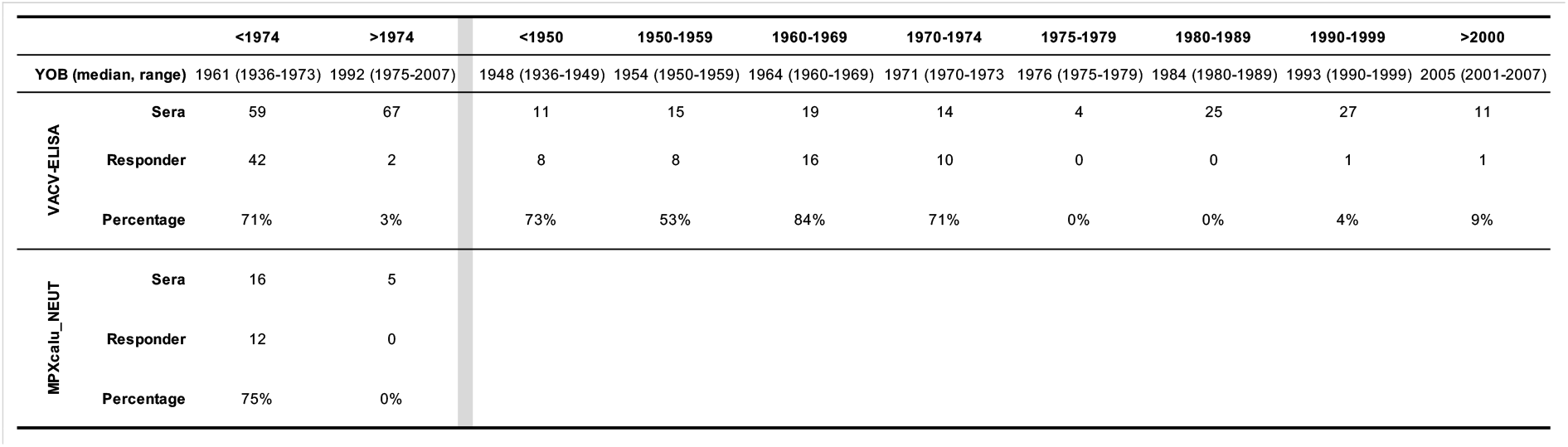
Overview of age-panel sera assessed for the presence of vaccinia virus (VACV)-reactive antibodies by ELISA and monkeypox virus (MPXV)-neutralizing antibodies by plaque reduction neutralization test (PRNT).

**Supplemental Table 2.** Overview of diagnostic panel sera assessed for the presence of vaccinia virus (VACV)-reactive antibodies by ELISA and monkeypox virus (MPXV)-neutralizing antibodies by plaque reduction neutralization test (PRNT).

|  |  | <1974 |  | >1974 |  |
| --- | --- | --- | --- | --- | --- |
|  |  | PCR-negative | PCR-positive | PCR-negative | PCR-positive |
| VACV_ELISA | Sera | 19 | 13 | 21 | 19 |
|  | Responder | 10 | 10 | 0 | 5 |
|  | Percentage | 53% | 77% | 0% | 26% |
| MPXcalu_NEUT | Sera | 11 | 12 | 5 | 7 |
|  | Responder | 7 | 10 | 0 | 5 |
|  | Percentage | 64% | 83% | 0% | 71% |

**Supplemental Table 3.** Overview of Imvanex panel sera assessed for the presence of vaccinia virus (VACV)-reactive antibodies by ELISA and modified vaccina virus Ankara (MVA) and monkeypox virus (MPXV)-neutralizing antibodies by plaque reduction neutralization test (PRNT).

|  |  | <1974 |  |  |  | >1974 |  |  |  |
| --- | --- | --- | --- | --- | --- | --- | --- | --- | --- |
|  |  | preVx | postVx1_2wk | postVx1_4wk | postVx2_4wk | preVx | postVx1_2wk | postVx1_4wk | postVx2_4wk |
| VACV_ELISA | Sera | 3 | 3 | 2 | 3 | 15 | 10 | 11 | 8 |
|  | Responder | 3 | 3 | 2 | 3 | 1 | 1 | 6 | 8 |
|  | Percentage | 100% | 100% | 100% | 100% | 7% | 10% | 55% | 100% |
| MVA_NEUT | Sera | 3 | 3 | 2 | 3 | 6 | 0 | 8 | 8 |
|  | Responder | 3 | 3 | 2 | 3 | 0 | 0 | 5 | 8 |
|  | Percentage | 100% | 100% | 100% | 100% | 0% | #DIV/0! | 63% | 100% |
| MPXcalu_NEUT | Sera | 3 | 3 | 2 | 3 | 6 | 0 | 8 | 8 |
|  | Responder | 3 | 3 | 2 | 3 | 1 | 0 | 5 | 5 |
|  | Percentage | 100% | 100% | 100% | 100% | 17% | #DIV/0! | 63% | 63% |

**Supplemental Table 4.** Overview of MVA-H5 panel sera assessed for the presence of vaccinia virus (VACV)-reactive antibodies by ELISA. (A) Responder rates in the 2 shot regimen. (B) Responder rates in the 1 shot regimen.

| **A** |  | 10<sup>8</sup> pfu MVA-H5 (2 shots) |  |  |  | 10<sup>7</sup> pfu MVA-H5 (2 shots) |  |  |  |
| --- | --- | --- | --- | --- | --- | --- | --- | --- | --- |
| VACV_ELISA |  | preVx | postVx1_4wk | postVx2_4wk | postVx3_4wk | preVx | postVx1_4wk | postVx2_4wk | postVx3_4wk |
|  | Sera | 0 | 7 | 2 | 9 | 6 | 5 | 10 | 3 |
|  | Responder | 0 | 1 | 2 | 9 | 0 | 0 | 6 | 3 |
|  | Percentage | #DIV/0! | 14% | 100% | 100% | 0% | 0% | 60% | 100% |

| **B** |  | 10<sup>8</sup> pfu MVA-H5 (1 shots) |  |  |  | 10<sup>7</sup> pfu MVA-H5 (1 shots) |  |  |  |
| --- | --- | --- | --- | --- | --- | --- | --- | --- | --- |
| VACV_ELISA |  | preVx | postVx1_4wk | postVx1_8wk | postVx2_4wk | preVx | postVx1_4wk | postVx1_8wk | postVx2_4wk |
|  | Sera | 9 | 2 | 8 | 1 | 5 | 8 | 2 | 9 |
|  | Responder | 0 | 0 | 6 | 1 | 0 | 4 | 1 | 9 |
|  | Percentage | 0% | 0% | 75% | 100% | 0% | 50% | 50% | 100% |

